# Examining the effect of smoking on suicidal ideation and attempts: A triangulation of epidemiological approaches

**DOI:** 10.1101/19007013

**Authors:** Ruth Harrison, Marcus R Munafò, George Davey Smith, Robyn E Wootton

## Abstract

**Background:** Previous literature has demonstrated a strong association between cigarette smoking and suicide-related behaviours, characterised as ideation, plans, attempts and suicide related death. This association has not previously been examined in a causal inference framework and has important implications for suicide prevention strategies.

**Aims:** We aimed to examine the evidence for an association between smoking behaviours (initiation, smoking status, heaviness, lifetime smoking) and suicidal thoughts or attempts by triangulating across observational and Mendelian randomisation (MR) analyses.

**Methods:** First, in the UK Biobank, we calculate observed associations between smoking behaviours and suicidal thoughts or attempts. Second, we used Mendelian randomisation (MR) to explore the relationship between smoking and suicide using genetic variants as instruments to reduce bias from residual confounding and reverse causation.

**Results:** Our observational analysis showed a relationship between smoking behaviour and suicidal behaviour, particularly between smoking initiation and suicidal attempts (OR = 2.07, 95% CI = 1.91 to 2.26, p<0.001). The MR analysis and single SNP analysis, however, did not support this. Despite past literature showing a positive dose-response relationship our results showed no clear evidence for a causal effect of smoking on suicidal behaviours.

**Conclusion:** This was the first MR study to explore the effect of smoking on suicidal behaviours. Our results suggest that, despite observed associations, there is no strong evidence for a causal effect of smoking behaviour on suicidal behaviour. Our evidence suggests that further research is needed into alternative risk factors for suicide which might make better intervention targets.

## Introduction

There are more than 800,000 deaths from suicide each year^1^, and for each death, there are 10-40 unsuccessful attempts^2^. The World Health Organisation (WHO) has recognised this significant public health problem and the need for comprehensive suicide prevention strategies, however at present there is limited evidence of sustained reductions in suicides rates^1^. Studies of the relationship between cigarette smoking and suicide have demonstrated a strong association between current cigarette smoking and suicide-related behaviours characterised as ideation, plans, attempts and suicide related death^3–20^. These associations have been shown to have positive dose–response relationships^7,8,14^ that remain after adjustment for potential confounding variables such as psychiatric symptoms^5^, familial risk factors^17^, socio-economic characteristics^6^ and alcohol consumption^18^. Smoking interventions such as imposing cigarette taxes and smoke free air policies are reported to be protective against suicide-related outcomes^9^.

A number of psychopathological and physiological hypotheses have been explored to determine whether this association is causal. Even after controlling for specific confounding variables, residual confounding could still be biasing our estimates^6^, for example as a result of social deprivation, lower levels of education and higher levels of impulsivity^8,11^. Another hypothesis is of reverse causation, that smokers with pre-existing mental illness may use nicotine to self-medicate^11^, and in addition, their illness could lead to reduced motivation for smoking cessation. However, there is an alternate possibility, that smoking acts as a causative agent^11^. A number of possible biological pathways have been explored^7^, including evidence that smoking lowers the levels of serotonin^15^ and monoamine oxidase A and B^23^. Reduced levels of these neurotransmitters are related to depressive episodes and low levels of serotonin are also linked to increased impulsivity^15^. Nicotine has been found to act as a potent activator of the hypothalamic-pituitary-adrenal (HPA) axis and this has been linked to suicidal behaviour^24^. There is evidence to suggest that chronic cigarette smoking has long-term neurocognitive effects which lead to increased impulsivity and difficulties with decision making due to impairments in cognitive flexibility^7,25^.

A clear understanding of the relationship between smoking and suicide remains to be established. A recent study demonstrated a causal link between smoking and risk of depression using a Mendelian Randomisation (MR) approach^26^. MR can be implemented as a type of instrumental variable analysis in which genetic variants known to be associated with the exposure (smoking) are used as an instrument to test for an effect on the outcome (suicidal ideation and suicide attempts). In this study, we apply MR techniques, using genetic variants identified in genome-wide association studies (GWAS), to the relationship between smoking and suicide. Previous observational research in this area could be biased residual confounding and reverse causation, and MR is one way to overcome these limitations^27^. In our analysis we looked at smoking initiation, smoking heaviness and lifetime smoking, using a genetic instrument that takes into account smoking status, duration, heaviness and cessation^26^.

## Methods

### Observational analysis

#### Sample

The UK Biobank is a research resource of health data collected on over 500,000 individuals from study centres located across the United Kingdom. Recruitment occurred between 2006 and 2010. Participants were aged from 39 to 70 years at recruitment (mean = 56.91 years, SD = 7.99 years) and 54% of the sample were female. Overall, 30% of the sample had ever smoked (8% current smokers and 22% former smokers). Further information is available elsewhere (https://www.ukbiobank.ac.uk/).

#### Measure of suicidal ideation

In the UK Biobank, participants were asked as part of a questionnaire on depressive symptoms “Over the last 2 weeks, how often have you been bothered by any of the following problems? Thoughts that you would be better off dead or of hurting yourself in some way” (field 20513). Participants could respond depending on frequency of thoughts either “not at all”, “several days”, “more than half the days” or “nearly every day”. We recoded these into a binary variable in which those who responded “not at all” were coded as 0 and everyone else was coded as 1. Individuals who responded “prefer not to answer” were coded as missing.

#### Measure of attempted suicide

In the UK Biobank, participants were first asked “Have you deliberately harmed yourself, whether or not you meant to end your life?” (field 20480). If they responded yes, then they were asked “Have you harmed yourself with the intention to end your life?” (field 20483). We used both of these measures to derive one binary measure of suicide attempt in which participants were given a score of 0 if they responded negatively to either question and a score of 1 if they responded affirmatively to both questions. Individuals who responded “prefer not to answer” were coded as missing.

#### Measure of smoking behaviours

Participants in the UK Biobank self-reported their smoking status (field 20116). All ever smokers were asked to report their average number of cigarettes per day (fields 3456 and 2887). If participants responded “do not know” or “prefer not to answer” they were coded as missing.

#### Statistical analysis

After restricting to individuals of European ancestry with genetic data available (to make this analysis comparable with subsequent analyses), 337,053 individuals remained. We looked at the effect of four smoking behaviours on suicidal ideation and attempts. These were: smoking status (ever v. never), smoking status (current v. former within ever smokers), cigarettes per day (within ever smokers) and lifetime smoking score. The latter is a combination of smoking duration, smoking cessation and smoking heaviness described in detail elsewhere^26^. The effect of each of these smoking behaviours on suicidal behaviour was estimated using logistic regression, controlling for age, sex and socio-economic position (SEP). All analyses were conducted using R^28^.

### Mendelian randomisation analysis using summary level data

#### Smoking instrument

This MR approach requires GWAS summary data from two independent samples. The GWAS of suicide attempt was conducted in the UK Biobank, therefore we were unable to use the lifetime smoking instrument because it was also constructed using the UK Biobank^26^. Instead we used the smoking initiation (ever v. never) GWAS conducted by the GSCAN consortium taking betas from the GWAS with UK Biobank removed^29^. 23andMe had to additionally be removed because they do not allow their full summary data to be released. GSCAN identified 378 conditionally independent genome-wide significant SNPs associated with smoking initiation which explain 4% of the phenotypic variance^29^.

#### Suicide GWAS

The GWAS of suicide attempts was conducted in the UK Biobank using the question and method outlined above with 337,199 participants of which 2,433 were cases^30^. The authors did not identify any genome-wide significant SNPs but the summary statistics can be used as an outcome sample in summary level MR.

#### Statistical analysis

All analysis was conducted using the TwoSampleMR package^31^ in R^28^. We used five different MR methods: inverse-variance weighted, MR Egger^32^, weighted median^33^, weighted mode^34^ and MR RAPS^35^. Each method makes different assumptions and therefore a consistent effect across multiple methods strengthens causal evidence^36^. If a SNP was unavailable in the outcome GWAS summary statistics, then proxy SNPs were searched for with a minimum LD r^2^ = 0.8 and palindromic SNPs were aligned if MAF<0.3. We also performed Rucker’s Q test of heterogeneity and the MR Egger intercept test to estimate potential directional pleiotropy^32^. Finally, we performed steiger filtering to test for possible reverse causation^37^.

### Mendelian randomisation analysis using individual level data

#### Genotyping

UK Biobank participants provided blood samples at initial assessment centre. Genotyping was performed using the Affymetrix UK BiLEVE Axiom array for 49,979 participants and using the Affymetrix UK Biobank Axiom® array for 438,398 participants. The two arrays share 95% coverage, but chip is adjusted for in all analyses because the UK BiLEVE sample is over represented for smokers. Imputation and initial quality control steps were performed by the Wellcome Trust Centre for Human Genetics resulting in over 90 million variants^38^.

Individuals were excluded if there were sex-mismatches between reported and chromosomal sex or aneuploidy (N=814). MRC Integrative Epidemiology Unit filtering restricted the sample to individuals of European ancestry based on the first four principal components of population structure^39^. After excluding individuals who had withdrawn consent, 463,033 of the participants remained^39^. We restricted our analysis to autosomes only and used stringent filtering thresholds for SNPs of MAF<0.01 and info>0.8.

#### Conducting the GWAS

Participants from the UK Biobank were randomly allocated to one of two split halves of the genetic data. We then generated lifetime smoking scores in sample 1 of these two samples and ran a GWAS using the UK Biobank pipeline^40^ following the exact method as described elsewhere^26^.

#### Genetic instrument

From the GWAS, genome-wide significant variants (p<5×10^−8^) were clumped for independence at 10000kb and r^2^<0.001.

#### Instrument validation

We tested the validity of this instrument by creating a polygenic score from these variants in the second sample from the UK Biobank. This was done using Plink and weighting each allele by the effect size identified in the GWAS of sample 1^41^. This therefore provides an independent replication sample to check how much of the variance is explained in lifetime smoking. If this significantly predicts lifetime smoking in the independent second sample, then this can be used as an instrument in the individual level MR analysis.

#### Statistical analysis

We conducted individual level MR using instrumental variable regressions run in R^28^ with the ivreg command from the AER package. The instrument was the polygenic score from the GWAS in sample 1. We controlled for age, sex, and 10 principal components of population structure in all analyses apart from when we ran the analysis separately in males and females. Then sex was removed as a covariate.

### Single SNP analysis

#### Statistical analysis

Best guess genotypes at the SNP rs1051730 were extracted using Plink^41^ in the UK Biobank full sample described above. This SNP in the gene cluster *CHRNA5-A3-B4* is known to be strongly associated with heaviness of smoking^42–45^. We tested using logistic regression whether the number of effect alleles (A) of this SNP were associated with risk of suicidal ideation and suicide attempts again using the measures described above. We controlled for age and sex in all analyses. The logistic regressions were run in each category of smoking status separately (ever, current, former and never smokers). A causal effect of smoking on suicidal behaviour would be characterised by an effect of rs1051730 in all categories of smoking status apart from never smoking which provides a negative control.

### Sensitivity analysis

#### Smoking and impulsivity

We wanted to ensure that any effects of smoking on suicide attempts were not the result of confounding from personality factors (e.g. impulsivity and risk taking) rather than direct effects of smoking. Therefore, we conducted a follow up analysis using bi-directional MR of smoking initiation on risk taking behaviour using summary level data. As the instrument for smoking initiation we used the 378 SNPs from the GSCAN consortium GWAS and effect sizes with UK Biobank removed^29^. For risk taking behaviour we used the SSGAC GWAS meta-analysed across multiple cohorts of European ancestry^46^ which identified 124 SNPs associated with risk tolerance. These analyses followed the method described above for summary level data.

### Ethical approval

UK Biobank has received ethics approval from the UK National Health Service’s National Research Ethics Service (ref 11/NW/0382) and this work is part of approved project 9142.

## Results

### Observational analysis

Of the 109,649 individuals who had responded to the question of suicidal ideation, 4,515 (4%) had had suicidal thoughts. Of the 110,035 individuals who had responded to the questions of suicide attempts and self-harm, 2,405 (2%) had ever attempted suicide. Using logistic regression, every smoking behaviour increased odds of suicidal behaviour with the greatest effect being of initiating smoking on odds of attempting suicide (Table 1).

**Table 1.**
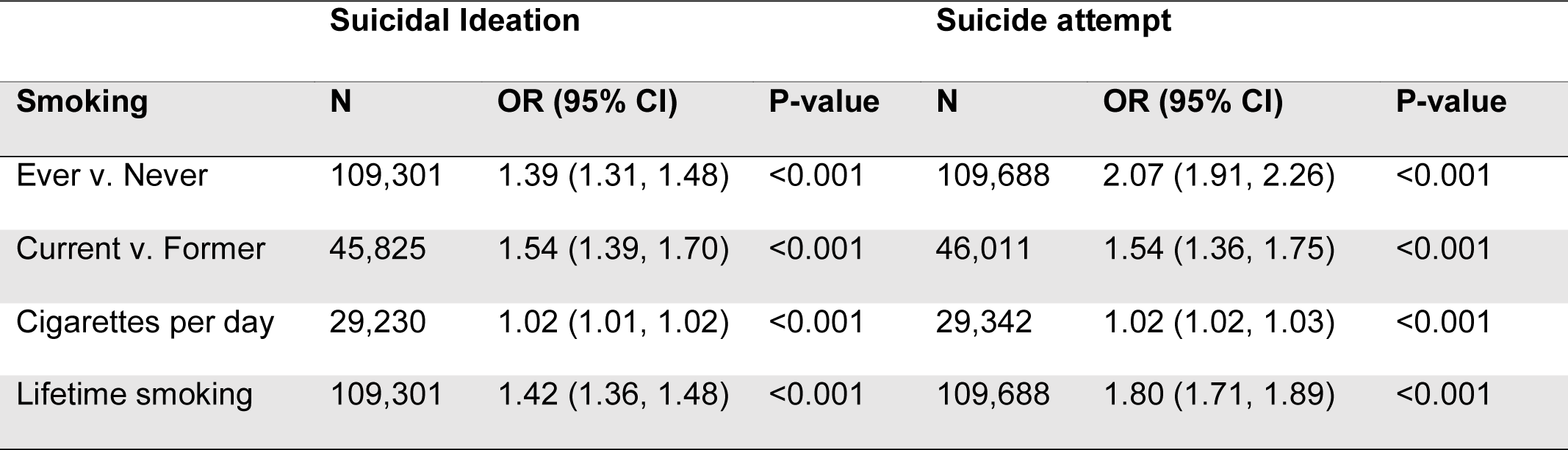
The observed association of smoking behaviour on suicidal ideation and suicide attempts controlling for age, sex and socio-economic position

### MR analysis with summary level data

Of the 378 conditionally independent SNPs associated with smoking initiation identified by the GSCAN consortium^29^, 321 were available in the GWAS summary data for suicide attempt^30^. We first performed the Rucker’s Q test of heterogeneity which did not provide evidence for heterogeneity (Supplementary Table S1). MR Egger analysis could not be conducted because the regression dilution I^2^_GX_ was below 0.6 (I^2^_GX unweighted_ = 0.07). All of the other four MR methods showed the same direction of effect with smoking initiation increasing the odds of attempting suicide (Table 2). The strongest evidence was from the IVW and MR RAPS methods. The evidence was weaker for the weighted median and weighted mode approaches which make different assumptions about the nature of pleiotropy. However, the MR Egger intercept and Rucker’s Q tests showed no clear evidence of bias from directional pleiotropy (Supplementary Table S2). Steiger filtering estimated that over half of the genetic instruments explained more variance in the outcome than the exposure suggesting that there might be reverse causation (Supplementary Table S3).

**Table 2.** Mendelian randomisation analyses using summary level data of smoking initiation on risk of suicide attempt.

| Method | N SNP | OR (95% CI) | P-value |
| --- | --- | --- | --- |
| Inverse-Variance Weighted | 321 | 2.42 (1.53, 3.83) | <0.001 |
| Weighted Median | 321 | 1.73 (0.85, 3.50) | 0.13 |
| Weighted Mode | 321 | 1.93 (0.24, 15.28) | 0.53 |
| MR RAPS | 321 | 2.86 (1.64, 4.98) | <0.001 |

### MR analysis using individual level data

To conduct an MR of lifetime smoking behaviour on risk of suicidal ideation and suicide attempt using individual level data, we first had to conduct a GWAS of suicide attempt in the one-half of the UK Biobank using a random split. We identified 19 independent genome-wide significant SNPs associated with lifetime smoking score. These were then extracted from the second half of the UK Biobank sample (with no sample overlap) and weighted by the effect size to create a polygenic score for each individual. The second half of the UK Biobank sample is 54% female with mean age 56.88 years (SD = 8.00 years). Mean lifetime smoking score in the second half of the sample was 0.342 (SD = 0.679). 4% of individuals had had suicidal thoughts (4% of females and 4% of males) and 2% of individuals had caused themselves harm with the aim to end their life (3% of females and 2% of males). We tested the association of lifetime smoking score and polygenic risk score on the baseline confounders of sex, age, socio-economic position (SEP), alcohol consumption and educational attainment (Supplementary Table S4). For all confounders, the association was attenuated for the polygenic risk score compared to the observed association.

We validated that the score predicts smoking behaviour by conducting a linear regression of polygenic score on lifetime smoking behaviour in the second half of the UK Biobank. It explained 0.171% (p<0.001) of the variance in lifetime smoking behaviour. Finally, we conducted the individual level MR analysis of lifetime smoking polygenic risk score on suicidal ideation and suicide attempt controlling for age, sex and 10 principal components of population structure (Table 3). There was no clear evidence for an effect of lifetime smoking on suicidal ideation or suicide attempt but a trend towards increased risk in both analyses (Table 3).

**Table 3.**
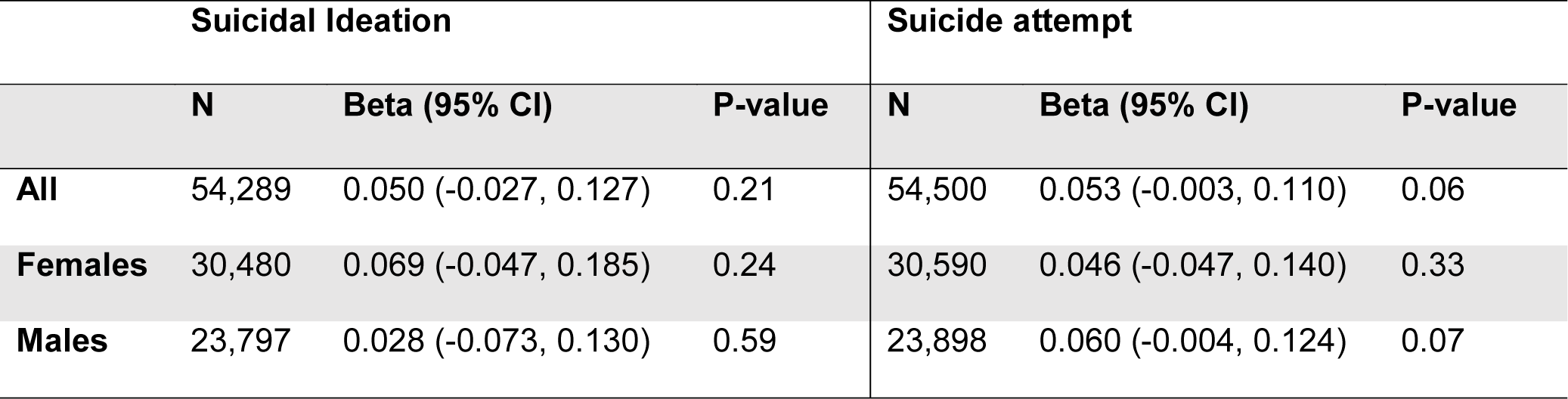
MR analysis of lifetime smoking on suicidal ideation and attempt using individual level data

### Single SNP analysis

Finally, we extracted values for the SNP rs1051730 (A/G) from individuals in the UK Biobank. We first confirmed that an increased number of effect alleles (A) were associated with increased smoking behaviour (Supplementary Table S5) where we observed the anticipated increase of ∼1 cigarette more per day per allele within ever smokers. We also showed that genotype at rs1051730 is not associated with smoking status or the baseline confounders of sex or alcohol consumption (Supplementary Table S5). However, there was some evidence to suggest that genotype at rs1051730 was associated with educational attainment and age (Supplementary Table S5). There was no clear evidence for an effect of rs1051730 genotype on suicidal behaviours controlling for age and sex (Figure 1). There was weak evidence to suggest that the number of rs1051730 effect alleles might *reduce* risk of suicide attempts with no effect in the never smokers and a small protective effect in the ever smokers (Figure 1). If smoking is increasing risk of suicidal behaviour, we would still expect to see no association in never smokers but the opposite effect within ever smokers.

**Figure 1.**
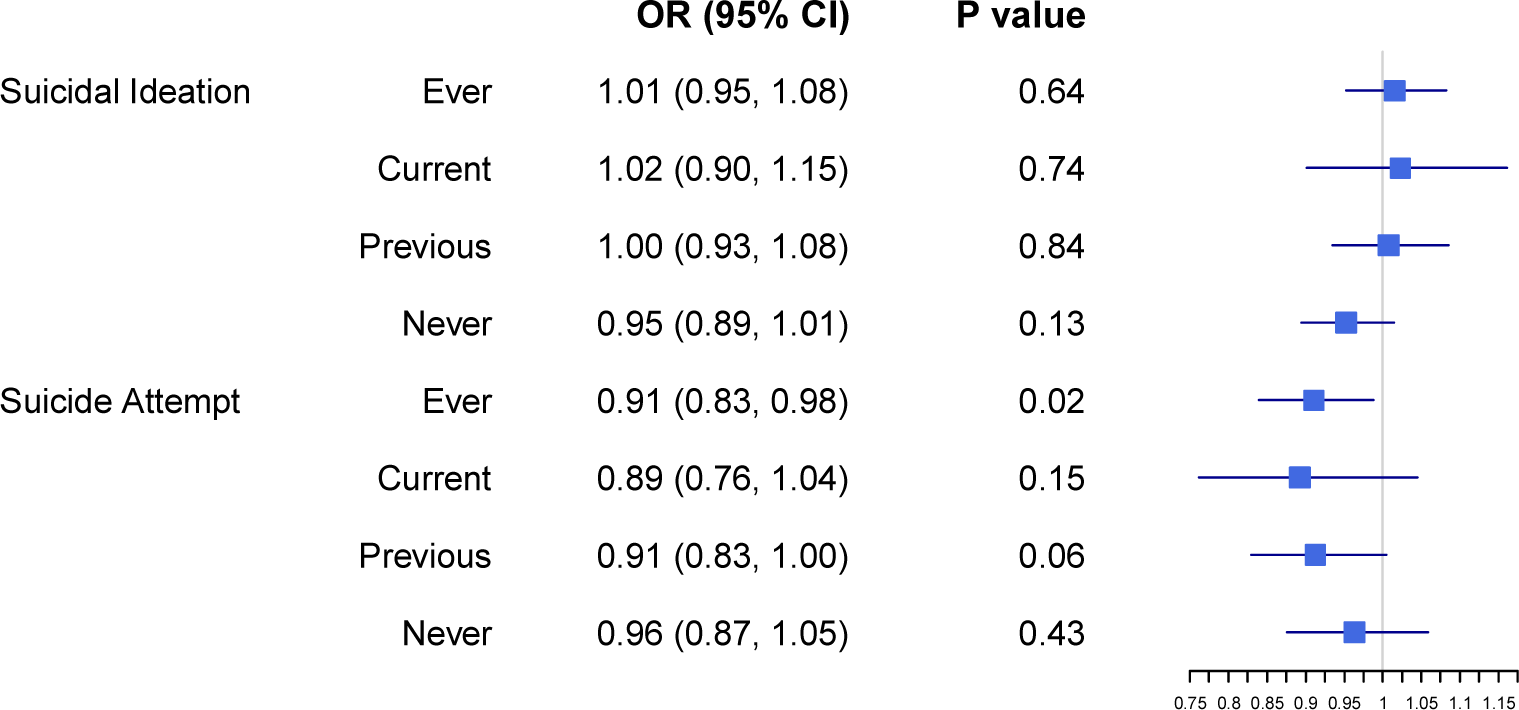
Logistic regression results of genotype at rs1051730 on odds of suicidal ideation and suicide attempt by smoking status.

### Smoking and impulsivity sensitivity analysis

We only saw evidence for smoking as a risk factor for suicide attempts when the instrument was smoking initiation. One trait associated with both smoking initiation and suicide attempts is impulsivity. Therefore, we conducted a bi-directional MR of smoking initiation and risk-taking using summary level data which have previously been shown to be genetically correlated^46^. There was strong evidence of a bi-directional causal relationship between smoking initiation and risk-taking behaviour suggesting that the smoking initiation SNPs might be capturing an underlying impulsivity phenotype (Supplementary Table S6) and this could explain why we only observed effects of smoking initiation on suicidal behaviour and not for the other smoking phenotypes. Rucker’s Q tests showed some evidence of heterogeneity (Supplementary Table S1) and the MR Egger intercept showed weak evidence of bias by directional pleiotropy (Supplementary Table S2).

## Discussion

The relationship between smoking and suicide-related behaviour is not clearly understood and has important clinical implications^1,2^. In this study, our observational analysis replicated previous observed associations between smoking behaviour and suicidal behaviour, particularly between smoking initiation and suicidal attempts. We went on to explore this association using a Mendelian Randomisation (MR) approach to understand if the association is causal. Overall, the MR analyses, including the single SNP analysis, did not support a causal interpretation. Therefore, despite past literature showing a positive dose-response relationship, our results do not support a causal effect of smoking on suicide.

Our results show evidence of a relationship between smoking initiation and suicidal ideation, but little evidence of an effect of lifetime smoking on suicidal ideation. Taken together with weak evidence for a protective effect in the single SNP analysis, these triangulated results overall suggest that there is only weak evidence for an effect of smoking on increased risk of suicidal behaviours.

### The potential role of impulsivity

The only evidence for smoking as a risk factor was when smoking initiation was the genetic instrument. However, our follow up analyses suggest this could be due to the instrument capturing underlying impulsivity. Smoking initiation is a complicated instrument with both behavioural and biological components. This behavioural component is likely to be related to impulsivity in part. This is supported by high genetic correlations between the two phenotypes^46^. As there is a known correlation between impulsivity and suicidal behaviours, we were mindful of this association. Therefore, to examine the impact of impulsivity on our results we undertook a bi-directional MR of smoking initiation and risk-taking. We showed strong evidence of a bi-directional causal relationship between smoking initiation and risk-taking behaviour suggesting that our results might be capturing impulsivity and not smoking. This is further supported by the fact that when we are using other instruments of smoking behaviour (e.g. lifetime smoking and smoking heaviness) we did not see any evidence for an effect.

Self-harm is the result of complex interactions of personality factors, including impulsivity, social factors and mental state^47^. It is well established that impulsivity is an important risk factor for suicidal attempts^48^. It has been shown that impulsivity increases with exposure to nicotine, returns to a normal levels with abstinence and increases with re-challenge after abstinence^49,50^. The association with suicidal attempts is not found in former smokers and this therefore supports the link between smoking and impulsivity^7,13,14,51^. This interaction between impulsivity, smoking initiation and suicidal attempts is complex and requires further research.

Recent theoretical models of suicide (i.e. integrated motivational volitional model^52^), all fit with the ‘ideation to action’ framework which posits that the development of suicidal ideation and progression from ideation to attempts are distinct processes with separate risk factors and explanations^53^. This has clearly important implications in clinical practice and risk management. Few studies have examined smoking within an ‘ideation to action’ framework. A cross-sectional study found suicide attempters were more likely than ideators to be current smokers^54^. However, in the National Comorbidity Survey, early onset nicotine dependence was prospectively associated with suicide plans but not attempts amongst those with ideation^12^. Our single SNP analysis would support the idea of suicidal ideation and attempts being separate processes, differentially affected by smoking heaviness and this differentiation is an important area which requires further research.

### Strengths and limitations

This study has many strengths, being the first to our knowledge to use the method of Mendelian randomisation to explore the association between smoking and suicide. We triangulated across multiple methods, multiple smoking behaviours and multiple suicidal behaviours to improve causal inference. However, the power of these analyses was limited by sample size. The single SNP analysis, designed to test the effect of smoking heaviness on suicidal behaviour, included small numbers of those experiencing suicidal thoughts and suicide attempts and was therefore underpowered. However, if anything the trend of association was in the opposite direction to what was hypothesised. It should also be noted that the sample size includes only non-fatal suicide attempts, and therefore our definition of suicide attempts is narrow. We were also limited by the UK Biobank questions asked for suicidal ideation and attempt. Suicidal attempt questions refer to lifetime attempts but suicidal ideation questions refer to symptoms. Another possible limitation is bias from reverse causation. As attempts were unsuccessful, a pathway from attempts to smoking initiation is possible. Furthermore, steiger filtering estimated that over half of the smoking initiation genetic instruments explained more variance in suicide attempts than smoking initiation.

### Conclusion

This was the first MR study to explore the effect of smoking on suicidal behaviours. Our results suggest that, despite observed associations, when we triangulate across multiple MR methods, there was little evidence for a causal effect of smoking behaviour on suicidal behaviour. It supports recent literature to suggest that suicidal ideation and attempts might need to be thought of as different processes and it has highlighted the complexity of unpicking the behavioural from biological components of smoking behaviours. Our evidence suggests that further research is needed into alternative risk factors for suicide which might make better intervention targets.

## Data Availability

This paper predominantly uses summary statistics from genome-wide association studies which are publicly available. The paper also uses data from the UK Biobank cohort which is available upon application.

## Acknowledgments

We are grateful to the participants of the UK Biobank and the individuals who contributed to each of the previous GWAS analyses conducted as well as all the research staff who worked on the data collection. This paper has used UK Biobank Application number 9142.

## Author Contributions

GDS and MRM conceived the study. REW conducted the analysis. RH reviewed the literature and provided clinical expertise. REW and RH drafted the initial manuscript. GDS and MRM advised and guided all stages of analysis. All authors assisted with interpretation, commented on drafts of the manuscript, and approved the final version. REW is the guarantor and attests that all listed authors meet authorship criteria and that no others meeting the criteria have been omitted.

